# Statin Eligibility Disparities with Transition from the Pooled Cohort Equations to the AHA PREVENT

**DOI:** 10.64898/2026.01.29.26345173

**Authors:** Xiaowei Yan, Qiwen Huang, Jiang Li, Hannah Husby, Powell Jose, Pragati Kenkare, Matthew Solomon, Fatima Rodriguez, Adrian M. Bacong

## Abstract

**Background:** The 2023 AHA PREVENT (Predicting Risk of Cardiovascular Disease Events) equations were expected to replace the 2013 ACC/AHA Pooled Cohort Equations (PCE) for estimating atherosclerotic cardiovascular disease (ASCVD) risk. The real-world implications of this transition on statin eligibility and disparity are unknown.

**Objectives:** To evaluate how transitioning from PCE to AHA PREVENT alters statin eligibility across risk thresholds and racial and ethnic subgroups.

**Design, Setting, and Participants:** Retrospective cohort analyses of adults aged 40–75 years without diabetes, LDL-C ≥190 mg/dL, or prior statin use from Sutter Health (2010–2024) and NHANES (2011–2020). Ten-year ASCVD risk was estimated using both equations. Weighted analyses were applied to NHANES data.

**Main Outcomes and Measures:** Statin eligibility at PREVENT-ASCVD thresholds (3%, 4%, 5%, 6%, 7.5%) compared with PCE ≥7.5%, and the proportion of individuals reclassified below PREVENT-ASCVD thresholds.

**Results:** Among 229,839 Sutter Health patients (mean age 53.7 years; 53.8% women), 22.3% had PCE risk ≥7.5%. Among individuals above PREVENT-ASCVD 5% threshold level (18.0% in the cohort) 94.7% also met PCE criteria. However, 6.3% (11,866) would lose eligibility, disproportionately affecting non-Hispanic Black adults (18.7%) compared with non-Hispanic Asian adults (3.3%) among individuals who were below PREVENT-ASCVD 5% threshold. In NHANES (n=3,226; representing 32.7 million adults), 9.4% overall and 21.7% of non-Hispanic Black adults with PCE ≥7.5% lost eligibility at PREVENT-ASCVD 5% threshold level.

**Conclusions:** Transitioning from PCE to PREVENT recalibrates statin eligibility and may disproportionately affect non-Hispanic Black adults. Disparity-focused monitoring is essential for clinical implementation of this new model.

## Introduction

Atherosclerotic cardiovascular disease (ASCVD) remains the leading cause of morbidity and mortality in the United States.^1^ Cholesterol lowering therapy (e.g., statins)remains a cornerstone of primary prevention.^2^ For over a decade, the 2013 ACC/AHA Pooled Cohort Equations (PCE) have served as the standard risk assessment tool for guideline-directed statin initiation.^3,4^ While widely adopted, the PCE have may systematically overestimate risk in contemporary populations and raises concerns related to using race as a predictor.^5–7^ These limitations prompted the development of the Predicting Risk of Cardiovascular Disease Events (PREVENT) equations.^8,9^ The PREVENT equations perform better compared to the PCE and have been validated in various study populations, including underrepresented race and ethnic groups..,^10–13^ The PREVENT-Total CVD equation has been incorporated in the newly released 2025 AHA/ACC/AANH hypertension guidelines and will impact clinical practices and public health in a profound way.^14^

Recent analyses applying the PREVENT equations to nationally representative data demonstrate that statin eligibility is highly sensitive to the choice of risk threshold. For example, at a PREVENT threshold of 4%, the number of adults identified as statin eligible is similar to that identified by the PCE 7.5% threshold.^14^ Additionally, a sizable proportion of adults previously eligible for statin therapy under the PCE framework may no longer meet eligibility under PREVENT.^15^ The clinical and public health implications of these shifts remain poorly understood.

This transition may raise critical questions regarding disparities in cardiovascular prevention. Historical underuse of statins has disproportionately affected women, racial and ethnic minority populations, and individuals with lower socioeconomic status.^16^ If adoption of PREVENT preferentially reclassifies certain demographic subgroups out of eligibility, existing inequities in prevention may be exacerbated. Conversely, PREVENT’s elimination of race as a predictor may mitigate structural biases inherent to the PCE. Whether these changes ultimately narrow or widen disparities in statin allocation is unknown.

Despite its methodological advances, the real-world impact of transitioning from PCE to PREVENT-based risk assessment has not been rigorously evaluated. Therefore, we use a diverse population from a large healthcare system, as well as national data, to assess the potential impact of transitioning from the PCE model to the PREVENT equations on statin recommendation across eligibility thresholds, especially among different race and ethnicity groups. Addressing these gaps is essential to ensure that the adoption of PREVENT improves, rather than undermines, disparity and effectiveness in primary prevention.

## Methods

### Study Setting

This is a retrospective cross-sectional study and approved by the institutional review board at Sutter Health, a large diverse integrated healthcare system in California. We identified the primary care population who are eligible for assessing cardiovascular risk using the same criteria as a published study, ^12^ with an extended study period (1/1/2010-12/31/2024) and those who had complete data for estimating 10-year PREVENT-ASCVD risk. To address primary ASCVD prevention using the PREVENT-ASCVD equation and align with eligibility for the PCE model, we further restricted patients to those who were 40-75 years old at the index date (the date we estimated the 10-year ASCVD risk), and excluded patients who had established diagnosis of diabetes, or very high cholesterol levels (low-density lipoprotein cholesterol (LDL-D)≥190 mg/dL). To assess generalizability, we applied the same criteria to National Health and Nutrition Examination Surveys (NHANES, January 2011 to March 2020) data to identify individuals eligible for primary prevention.

The PREVENT-ASCVD 10-year risk score and the PCE risk score were calculated for each individual. In order to find a threshold for the PREVENT-ASCVD score that was similar to the 7.5% threshold based on the PCE model for statin initiation,^3^ we dichotomized the cohort using 7.5% based on the PCE risk score, where greater or equal to 7.5% was the “eligible for statin” group, and less than 7.5% was the “not eligible” group. The optimal threshold for PREVENT-ASCVD risk was determined by optimizing the Youden Index (i.e. sensitivity+ specificity-1) by fitting a logistic regression model that dichotomized PCE status as the outcome, and PREVENT ASCVD risk as a sole predictor. Other proposed thresholds (e.g., 3.0%, 4.0%) were tested as potential candidates for statin initiation cutoff score based on the PREVENT-ASCVD risk.^17^

In order to understand how changing the PREVENT-ASCVD risk threshold impacts patients’ eligibility for statins, the percentage of patients who met eligibility for statins based on PCE risk were calculated among these were equal or above the PREVENT-ASCVD risk threshold (i.e., analogous to true positive rate), and separately for below the risk threshold (i.e., analogous to false negative rate). The latter implied that patients are no longer eligible for statins if transition to the PREVENT-ASCVD risk score. We calculated the percentage for two PCE risk levels: 5%-7.4% (borderline risk), and ≥ 7.5% (intermediate risk), varying PREVENT-ASCVD thresholds.

Additionally, the absolute risk reduction (ARR) for patients who fell in each PCE category was calculated based on the assumption of 40% reduction in LDL-C with statin using an established published formula published. ^17,18^ We did this for the overall cohort, and by race and ethnic group was Analyses were repeated for the NHANES data, with application of sampling weights to each wave of the survey to obtain population representative numbers. The summary statistics for each variable of the final cohort across all waves included in this study were the average across these waves.

## Results

### Sutter Health Cohort

As shown in Figure 1, among 1,713,016 adult primary care patients, 425,661 patients were 30-79 years old and had complete data to calculate PREVENT-ASCVD and PCE. In total, 195,822 patients were further excluded due to having a baseline statin (n=93,959), or having diabetes (n=83,830) or high LDL-C (n=6,065) or age 30-39 years or >75 years of age (n=94,770), and the final analytical cohort included 229,839 patients with age ranging from 40-75 years old.

**Figure 1.** Cohort Flow Diagram

The optimal threshold was 6% by optimizing Youden Index in the relationship between PREVENT-ASCVD risk score and PCE threshold for statin initiation. Therefore, 6% as a threshold was added to the multiple hypothetical cutoff points (i.e., 3%, 4%, 5%, 6%, and 7.5%) in the analysis.

More than 22% (n=51,254) of patients had equal or above 7.5% risk based on PCE, 10.9% (n=25,053) had borderline risk (i.e., PCE 5%-7.4%) based on PCE, and 66.8% (n=153,522) had low risk of ASCVD (i.e., PCE<5%). Based on the PREVENT-ASCVD risk score, 18.0% (n=41,494) were equal or above 5%, 17.3% (n=39,837) had risk between 3% to 5%, and 64.6% (n=148,508) were under 3% risk (Table 1).

**Table 1.**
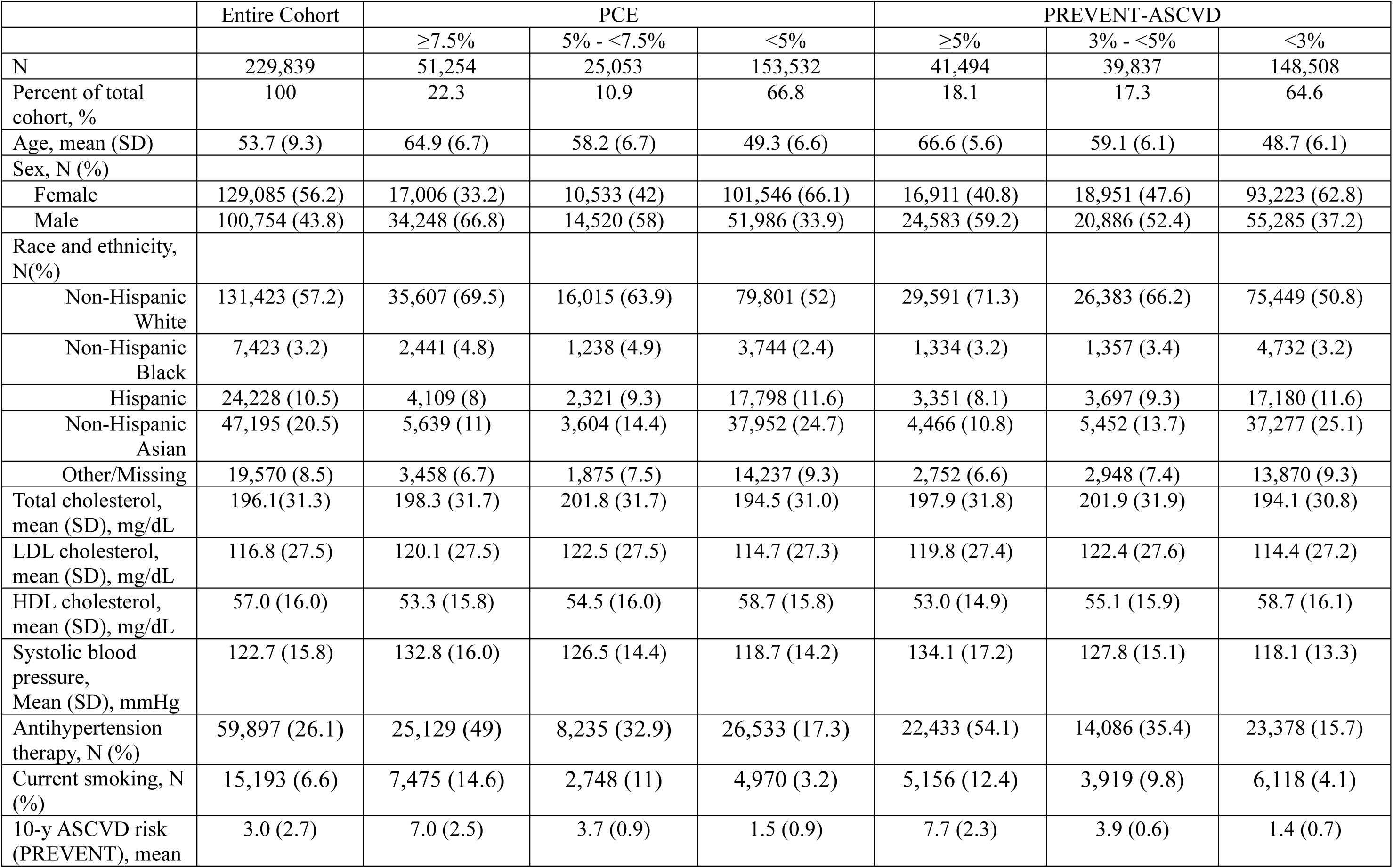
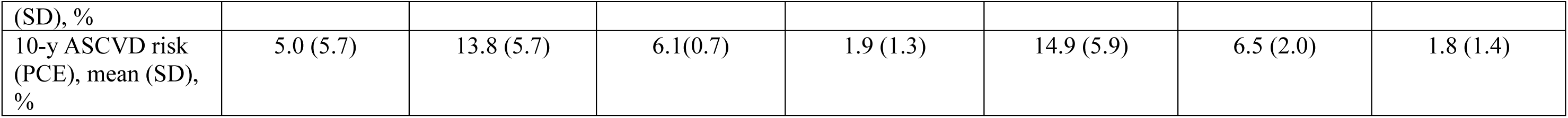
Patient Characteristics of Sutter Study Cohort (total cohort size=229,839)

As shown in the Supplemental Table S1 for the overall Sutter cohort, among 81,331 (35.4%) patients whose PREVENT-ASCVD risk were equal or above 3.0%, a low threshold, 62.5% fell in PCE ≥ 7.5% level and therefore were eligible to initiate moderate intensity statin, and 24.9% fell in the borderline level (5%-<7.5%). Among patients whose PREVENT-ASCVD score was ≥5% (18% out of the entire cohort) (Figure 2a, supplemental table S1), 94.7% would also meet PCE statin eligibility criterion. Almost all patients (99.4%) would be eligible for statin based on current guidelines if PREVENT-ASCVD risk threshold was 7.5%, which is the current risk cutoff for PCE (Supplemental Figure S1a).

**Figure 2.**
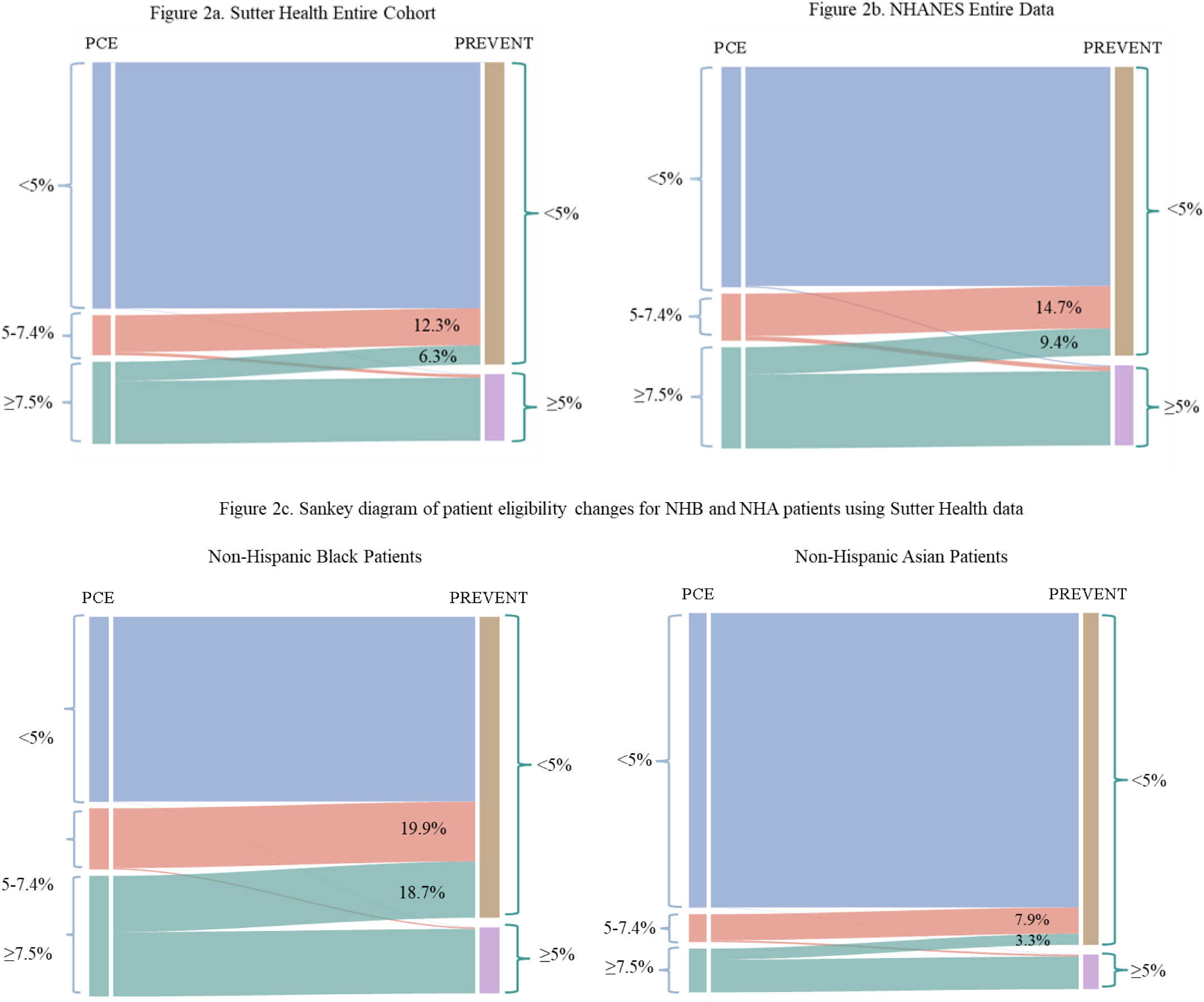
Sankey diagrams of patient eligibility changes between risk models

Separately, we evaluated eligibility change among patients who were below a PREVENT-ASCVD threshold thus no longer eligible for statin and how the impact varied by different thresholds (Supplemental Figure S2a). At a 3% threshold level, 64.6% of all Sutter cohort were below that threshold, among which, about 0.3% were above PCE ≥ 7.5%, and 3.2% were at PCE borderline level (i.e., 5% -<7.5%) (Supplemental Figure S2a). As the threshold increased to 5%, among patients who were below that threshold (81.9% out of entire Sutter cohort), a significant number of patients (n=11,866, 6.3%) would have met statin eligibility criteria based on PCE risk, translating to about 1.3% absolute risk reduction if those patients take statin. At 7.5% threshold level, as high as 16% (n=13,380) out of 212,388 who would have met statin eligibility based on PCE risk were no longer eligible for statins, corresponding to 1.6% absolute risk reduction which can be missed (Supplemental Figure S2a, supplemental table S1).

This impact varied substantially across racial and ethnic groups. As shown in supplemental Figure S1a and supplemental Table S1, among patients who were above PREVENT-ASCVD 5.0% threshold level, the percentage of patients who retained statin eligibility based on PCE risk varied from 93.4% (Hispanic patients) to 97.6% for non-Hispanic Black (NHB) patients, resulting in a relatively consistent 2% risk reduction. However, among patients who were below that threshold, 3.3% (non-Hispanic Asian, or NHA) to as high as 18.7% NHB patients would no longer be eligible for statins based on this threshold (Figure 2c), translating to about a 1% risk reduction that may be missed (Supplemental Table S2). Across all thresholds, NHB patients had the highest proportion who would lose statin eligibility, followed by Non-Hispanic White (NHW) patients, and was more than 3 times higher than NHA patients. At 7.5% PREVENT-ASCVD threshold level, more than quarter of NHB patients (27.2%) would lose eligibility for statins (Supplemental Figure S2a).

### NHANES Data

In total 15,043 participants, representing 131,696,623 adults who were 40-75 years old, were included in the analysis. After excluding 5,360 people already on statins/missing statin information, 5,609 with existing CVD/missing CVD information, 2,870 with diabetes, and 189 with elevated LDL-C, 3,226 participants, representing 32,660,990 eligible adults were included in the analysis (Table 2).

**Table 2.**
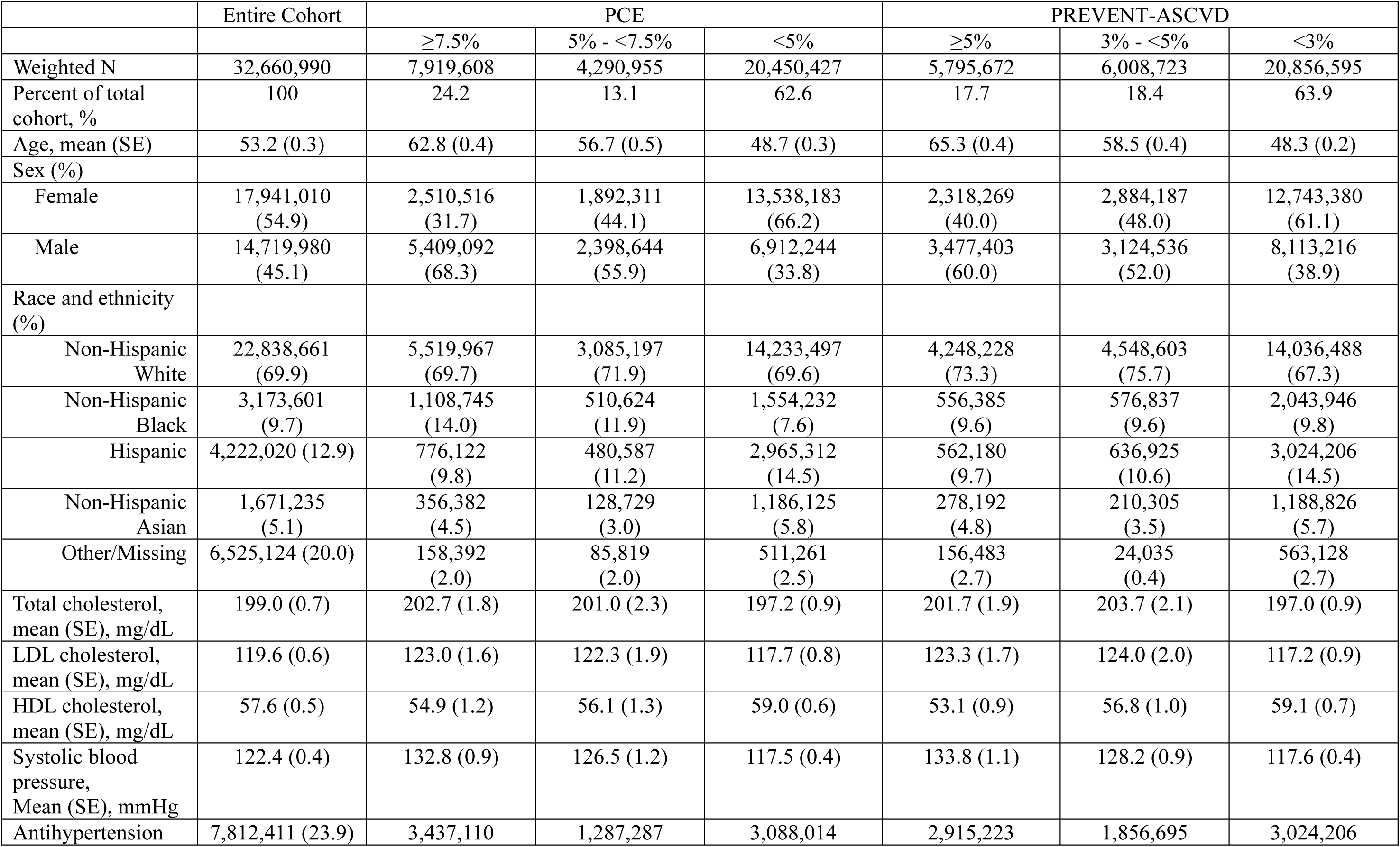
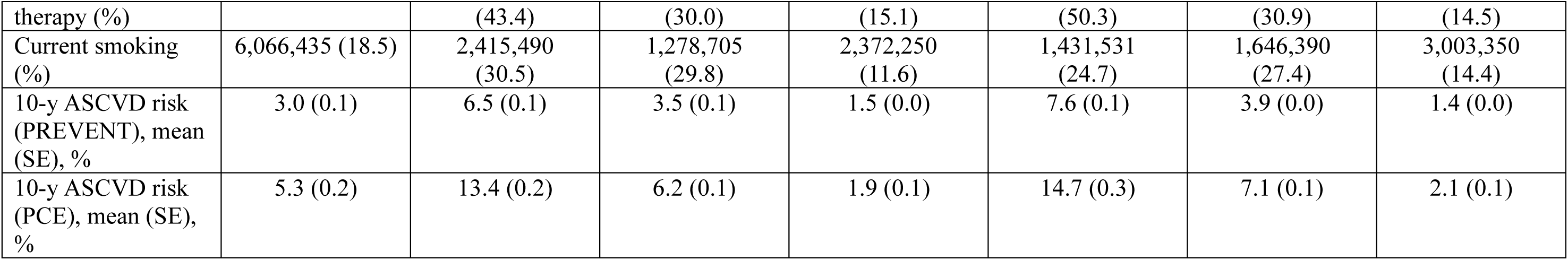
Survey Weighted Patient characteristics from U.S. population who are eligible for statins for primary prevention, National Health and Nutrition Examination Survey (NHANES), Weighted N = 32,660,990.

As shown in Table 1 and Table 2, the average age was similar between NHANES sample and Sutter cohort (53.2±0.3 for NHANES, 53.7±9.3 for Sutter cohort), while NHANES sample had significant lower proportion of NHA patients (5.1% in NHAHES vs. 10.5% in Sutter), higher proportion of NHB patients (9.7% in NHAHES vs. 3.2% in Sutter) or in missing/other racial and ethnic category (20.0% in NHAHES vs. 8.5% in Sutter). Total cholesterol and systolic blood pressure were similar between both samples. A slightly smaller proportion of people in the NHANES sample were on antihypertension medication (23.9%) when compared to the Sutter cohort (26.1%), while a larger proportion of people in the NHANES reported current smoking (18.5%) when compared to the Sutter cohort (6.6%).

In total 24.2% (weighted n=7,919,608) patients were categorized as moderate risk (i.e., ≥7.5%) for ASCVD based on PCE and 13.1% (weighted n=4,290,955) were at borderline risk for ASCVD. For PREVENT-ASCVD, 17.7% (weighted n=5,795,672) had PREVENT-ASCVD risk ≥5%, while 18.4% (n=6,008,723) had a risk score from 3%-4.9% (Table 2).

Patterns of changes in eligibility were similar to the Sutter cohort (Supplemental Figure S1b, Supplemental Table S3). When using a 7.5% threshold for PREVENT-ASCVD, 96.9% of people among PREVENT-ASCVD Score ≥ 7.5% also had a PCE score ≥ 7.5%. When using a PREVENT-ASCVD threshold at 5%, nearly 93% of people had a PCE ≥ 7.5%.

However, when examining people who lost eligibility (Figure 2b, Supplemental Table S4, Supplemental Figure S2b), using a 7.5% threshold (i.e. PREVENT-ASCVD <7.5%) resulted in 19% of people losing eligibility (i.e., PREVENT-ASCVD <7.5% and PCE ≥ 7.5%). At a 5% threshold, the percentage of people who lost eligibility was 9.4% (Figure 2b).

There were notable disparities in losing eligibility by race (Figure 3 NHANES, Supplemental Table S4). At a 7.5% threshold, 29.5% of NHB individuals with a PCE ≥ 7.5% would have lost eligibility. The other race and ethnic groups had notably lower percentages of people with a PCE ≥ 7.5% who would lose statin therapy eligibility (e.g., NHW = 18.8%, Hispanic = 13.9%, NHA = 16.8%). When the threshold is lowered to 5%, the proportion of people who would lose eligibility also decreased. Only 8.8% of NHW individuals would still lose eligibility for statin therapy if the statin threshold was 5%, followed by 6.8% of NHA and 6.4% of Hispanic individuals. However, 21.7% of NHB individuals with a PCE ≥ 7.5% would lose eligibility for statin therapy if a 5% threshold was used. In the sensitivity analyses where we removed the use of a “Black specific” equation for the PCE, only 8.6% of NHB individuals would lose eligibility (Figure 3 NHANES).

**Figure 3.**
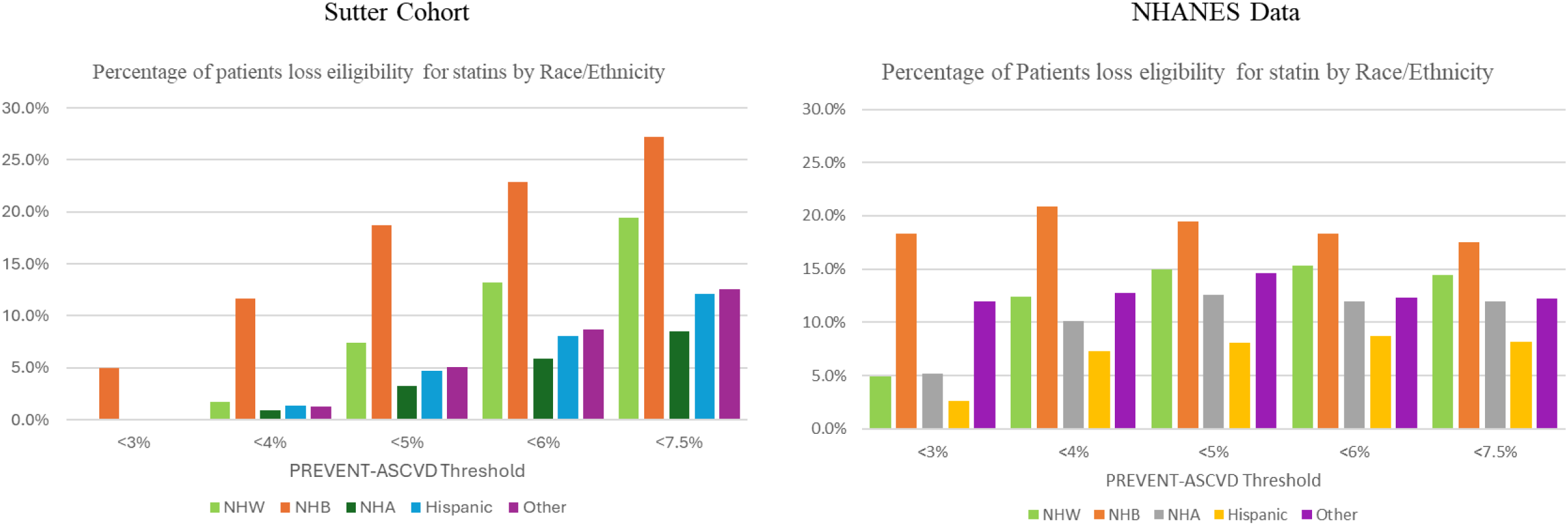
Percentage of patients losing original eligibility for statins among those who were below PREVENT-ASCVD threshold at different threshold levels, by race and ethnicity

## Discussion

In a recent publication, Khan et. al. recommended 5% risk as the statin eligibility threshold based on the PREVENT-ASCVD risk score,^17^ which is comparable to the 7.5% threshold based on PCE, yielding almost an equivalent number of individuals who meet eligibility for statins for primary prevention. We further examined the percentage of patients who might lose eligibility when the risk estimator transitions from PCE to PREVENT-ASCVD and at varying thresholds. In our study, about 6% of patients might lose eligibility at 5% risk as the threshold, equivalent to 11,865 patients. However, as high as 18.7% of NHB patients are no longer eligible for statins based on the new model at 5% risk threshold, leading to a disproportionally high (9.6%, i.e., 1138/11865) percentage of NHB patients among patients losing eligibility, compared to the proportion of NHB in the study cohort (i.e., 3.2%). Our results were replicated using national data, showing that as high as 21.7% of NHB patients no longer meet the statin initiation criteria if transitioning to the new risk estimator.

External validations have consistently concluded that PREVENT equations outperformed PCE, and once translated to clinical guidelines, clinicians would have confidence to use it as a shared-decision point to initiate conversations on CVD prevention ^11–13,19^. The ACC/AHA joint committee recently published hypertension clinical guidelines,^14^ which is the first clinical guideline officially adopting the PREVENT risk model and recommended 7.5% as threshold based on PREVENT-CVD risk to initiate antihypertension medication. Once clinically sensible thresholds for treatment initiation or titration are determined, this new risk estimator would be widely adopted in clinical practices. Recently 5% was suggested as the cholesterol lowering therapy (e.g., statins) initiation threshold based on PREVENT-ASCVD model.^17^ Our data and NHANES data have confirmed approximately 95% of patients who meet this new threshold also meet statin initiation guidelines based on PCE (i.e. 7.5% risk) overall and across all race and ethnic groups.

However, caution is warranted when eligibility criteria for primary prevention change. PREVENT equations have significantly improved prediction, adjusting the overestimation, and should yield a fairer treatment decision. However, improved accuracy of the new estimator requires an appropriate threshold to balance benefits and harms. First, some patients might lose eligibility for initiating cholesterol lowering therapy. About 11,866 patients (i.e., 6.3%) out of 188,345 at Sutter (or 9.4% in NHANES data) met the previous PCE ≥7.5% threshold but not this PREVENT-ASCVD ≥5% threshold, recommended by Khan et. al, ^17^ implying they might lose eligibility to cholesterol lowering therapy. As a result, it misses the opportunity of 1.2% absolute risk reduction if they were on statins. Second, how to choose threshold. An analytical threshold derived based on Sutter cohort or NHANES samples was 6%, which minimized the size of patients who were false positive or false negative based on PREVENT-ASCVD estimator compared to 7.5% threshold PCE estimator. Using this threshold, 10-14% among those whose PREVENT-ASCVD risk less than 6% would have been eligible for statins in PCE model. However, our study further shows that varying the threshold does not result in reduced disparities in eligibility change. Finally, the impact of this transition is not uniform across racial and ethnic groups. A significantly higher percentage of NHB patients (18.7% vs. 3.3% for NHA from Sutter data, and 19.5% vs. 8.1% for from NHANES data, Figure 3) fall into the category who might lose eligibility for statins. This might be partially explained by differential model performance for the new (i.e., PREVENT) and current race-specific risk estimator (i.e., PCE). The calibration analysis in the published study showed PCE overestimated risk by almost 50%, and PREVENT-ASCVD slighted underestimated for NHB, whereas among NHA, PCE overestimated the risk by ∼70%, and PREVENT-ASCVD slightly overestimated by 14%.^12^ Therefore, the risk reallocation can be the result of the different calibration performance between PCE and PREVENT-ASCVD, and this difference was manifested the most among the NHB group than any other racial and ethnic groups. A sensitivity analysis was conducted to use an identical pooled cohort equation (i.e., the PCE that is used for White and other race groups) to estimate risk for NHB patients,^20^ and the result showed that the percentage switching from eligible to noneligible dropped from 18.7% to 5.6% for NHB patients (Supplemental Table S2). This suggests that disparity should be considered as a part of a quality metric when evaluating a risk estimator, a suboptimal estimator can lead to potentially wider disparities.

Clinicians may be more cautious regarding what clinical recommendations should be given to their patients during this risk model transitional period, especially when recommendation based on old model is inconsistent with the new recommendations. Tools for further risk stratification might be needed for decisions on cholesterol treatment, such as coronary artery calcium (CAC) score, lipoprotein (a) (Lp(a)) testing, family history of premature CVD, or other identified CVD risk enhancement. Cost must also be considered as one critical contributor in making recommendations.

### Limitations

This study has some notable limitations. First, cholesterol-lowering therapy guidelines incorporating PREVENT have not yet been published, and thus the definitive treatment threshold remains uncertain. We evaluated hypothesized thresholds and highlighted both potential benefits and concerns, particularly regarding differential racial/ethnic impacts during the transition period. Second, we did not assess other PREVENT-based equations, such as PREVENT-CVD, which has already been incorporated into hypertension guidelines. Third, ASCVD risk estimation is only one component of decision-making, additional risk-enhancing factors must be considered, and clinical decisions should ultimately be guided by shared decision-making between patients and clinicians. Fourth, we assume good adherence of statins once prescribed in estimating absolute risk reduction, which is not always true. Low adherence to statins was commonly reported in the literature, ^21–23^ thus the observed absolute risk reduction might be much lower. Finally, we did not examine downstream associations with clinical outcomes such as ASCVD events or LDL-C trajectories. Longitudinal studies are needed to evaluate the real-world impact of transitioning from PCE to PREVENT.

## Conclusions

PREVENT equations are poised to replace PCE in forthcoming cardiovascular prevention guidelines. While PREVENT demonstrates improved calibration and equity across most racial and ethnic groups, our findings highlight that non-Hispanic Black patients may be disproportionately affected by changes in eligibility. Transparent communication of these subgroup effects during guideline adoption will be critical to ensure equitable implementation and maintain trust in preventive cardiovascular care.

## Data Availability

This study utilized Electronic Health Record data, under the institute policy, data will not be shared due to patient PHI contained in the data.

